# Breaking down barriers in quality medical laboratory services: insights from a qualitative study of health managers in Ghana

**DOI:** 10.64898/2025.12.09.25341931

**Authors:** Elizabeth Sorvor, Mawuli Dzodzomenyo, Justice Nonvignon, Genevieve Cecilia Aryeetey

## Abstract

**Background:** Quality medical laboratory services are vital for accurate diagnosis, effective treatment and public health surveillance. However, various barriers in low- and middle-income countries, such as Ghana, hinder the consistent provision of high-quality laboratory services. Understanding these barriers is crucial for developing targeted interventions and enhancing medical laboratories to achieve improved health outcomes. This study examines the barriers to delivering high-quality medical laboratory services in Ghana.

**Methods:** A qualitative approach was employed to understand and explore barriers associated with providing quality medical laboratory services. The study was conducted in the Kumasi Metropolis, Ashanti Region, Ghana, from March 8th to November 26th, 2021. Face-to-face interviews were conducted with seven (7) health managers and thirteen (13) laboratory managers across various levels and types of healthcare facilities (Government, Christian Health Association of Ghana (CHAG), and Private). Inductive and deductive multistage thematic analysis was used to interpret the data.

**Results:** The thematic analysis identified key themes shared among all participants, highlighting the main barriers to delivering quality medical laboratories as “Laboratory Financing,” “Quality Assurance,” “Human Resource Capacity,” “Laboratory workspace and facilities **“,** and “Policy and Regulation.”

**Conclusion:** Financial constraints and the limited control exercised by medical laboratories within the healthcare sector constitute significant barriers to delivering quality medical laboratory services.

## Introduction

Medical laboratory services are crucial for an effective health system, providing vital information that supports clinical decision-making, treatment, and disease surveillance [1–3]. It is estimated that over 70% of clinical decisions are influenced by laboratory testing, emphasizing the vital role that medical laboratory services play in improving patient outcomes and the overall performance of health systems [4]. It has been suggested that priority should be given to medical laboratories to strengthen and support health systems in achieving higher quality care [5–7].

Given the essential role of medical laboratories in producing clinical data, it is concerning that they have been largely ignored [8]. Evidence indicates that the medical laboratory is among the top three functions for enhancing key performance indicators (KPIs), the most prominent health informatics infrastructure, and an underutilized source of greater value in healthcare [8]. Consequently, the sidelining of laboratories in healthcare often leads to an unsustainable approach to health [9].

In low- and middle-income countries, challenges such as infrastructure, human resources, quality management, and systemic issues hamper the delivery of high-quality laboratory services [2,10–12]. Overcoming these barriers and tapping into the value that laboratories contribute can significantly improve the efficiency and quality of integrated healthcare [8]. Recognizing the vital roles of medical laboratories in delivering high-quality healthcare requires shifting from viewing them as mere commodities to appreciating them as essential components of the healthcare system [8].

In Ghana, medical laboratories are key to addressing the high burden of infectious diseases such as Tuberculosis, HIV, and malaria, as well as the rising prevalence of non-communicable diseases (NCDs). The increasing global health threats, including antimicrobial resistance and emerging epidemics, further highlight the necessity for high-quality laboratory services. Despite their importance, reports indicate that Ghana’s laboratory services at all levels are of poor quality, with unclear oversight and fragmented quality strategies and techniques that have little impact on patient experiences and health outcomes [13,14].

While research has examined certain aspects of medical laboratory service delivery in Ghana [14–17], significant gaps remain in the comprehensive evaluation of the various barriers affecting the quality of laboratory services across different healthcare settings. Gaining insights into these barriers is essential for implementing targeted interventions and enhancing the effectiveness of medical laboratories to achieve better health outcomes. This study investigates the barriers impeding the delivery of quality medical laboratory services and provides empirical evidence to address critical issues that must be prioritized to strengthen Ghana’s medical laboratory systems for improved healthcare.

## Materials and methods

### Study design and setting

The study employed a cross-sectional design with a qualitative approach. The study was conducted in the Kumasi Metropolis, Ashanti Region, Ghana. Its unique central position makes it accessible from all corners of the country. It is the second-largest city in the country and the administrative capital of the Ashanti region. Kumasi has the highest number of healthcare facilities, representing 38% of the total number of facilities in the Ashanti region. There are about two hundred and fifteen (215) health facilities in the metropolis, made up of hospitals, health centers, standalone laboratories, and clinics. The majority (87%) of health facilities in the metropolis are privately owned. Kumasi was chosen for the study because it serves as a major referral center for adjoining communities and provides a blend of all types of healthcare facilities, and thus is expected to provide high-quality healthcare for the population.

### Sampling and data collection

Face-to-face interviews were conducted with seven (7) health managers and thirteen (13) Laboratory managers in the healthcare delivery chain across different types of healthcare facilities, namely Private, Government, and Christian Health Association of Ghana (CHAG). These managers were recruited using stratified purposive sampling based on the crucial role they play in providing healthcare. The participants include the Director of Health Services, the Director of Laboratory Services, the Medical Director of Health Services, Hospital administrators, Medical superintendents, and Laboratory managers. All interviews were conducted in English, tape-recorded, and transcribed; the average interview time was 45 minutes. Interviews were conducted from 8^th^ March to 26^th^ November 2021.

### Data analysis

Thematic analysis was used to analyse the data. Interviews were transcribed verbatim into Microsoft Office. The transcript document was read multiple times to become familiar with all aspects of the data, and codes were assigned to specific pieces of data using NVivo 11 software. Codes were sorted and converted into themes and subthemes using deductive and inductive approaches. The constructed themes were based on common phrases. To convey participants’ impressions and enhance the explanatory power of the findings, relevant statements are quoted verbatim.

### Ethical consideration

This study is part of the PhD research work of the first author and received ethical approval from the Ghana Health Service Ethics Review Committee (Reference #: GHS-ERC: 014/11/20) and Komfo Anokye Teaching Hospital Institutional Review Board (IRB) (Reference #: KATH IRB/AP/155/20). Consent was obtained individually before data collection, and participants were given sufficient time to read and sign a consent form. The privacy and confidentiality of study participants were ensured by assigning codes rather than using their names.

## Results

### Characteristics of the Study Participants

The study involved 20 health managers, with 80% being male and 20% female. Most participants (50%) were employed in government hospitals, and the predominant profession (67%) was that of a Medical Laboratory Scientist (Table 1).

**Table 1.** Sociodemographic Characteristics of Participants

| Variable | Category | Frequency |
| --- | --- | --- |
| Gender | Male | 16 |
|  | Female | 4 |
| Facility type | CHAG | 5 |
|  | Government | 10 |
|  | Private | 5 |
| Profession | Medical doctor | 4 |
|  | Administrator | 2 |
|  | Medical laboratory scientist | 14 |
| Work experience | 3-10 years | 9 |
|  | 11-18 years | 7 |
|  | 19-26 years | 4 |

### Interview findings

The main theme and subthemes that emerged from the interviews, regarding the barriers affecting the provision of quality medical laboratory services, are discussed in the following section.

#### Laboratory financing

Key stakeholders recognized the essential role that financing plays in ensuring the quality of medical laboratories. The study identified inadequate budget allocation and the lack of a dedicated laboratory fund as two major challenges in this area.

##### Allocation of funds

Most health managers believed that although the laboratory generates funds from its service provision, these funds are not accessible for the inputs and reagents necessary for the smooth operation of the laboratories. The Director of Laboratory Services and a Laboratory Manager explain.

> *“The revenue generated by facility laboratories is often merged with the facility’s general funding, which means that laboratory managers do not have control over how much they can spend on equipment and reagents, despite being aware that their services have generated a significant amount of funding”* (**Regional Director of Laboratory Services**).
>
> *“There are instances where essential items are unavailable due to budget constraints, hence financing for laboratory operations is the primary obstacle to providing top-notch medical laboratory services” (**CHAG, Laboratory Manager**).*

##### Creation of laboratory fund

Some health managers believed that setting aside some funds solely for laboratory services would help provide uninterrupted service and potentially improve the quality of laboratory services.

> *“If the money we make is kept in a separate account, then we will always be able to pay the suppliers, and therefore, the reagent will not run out, but at the moment, the hen that is laying the golden eggs is not fed to lay the egg, so it is a problem.”* (**Tertiary Hospital, Director of Laboratory Services**).
>
> *“We must establish a fund for laboratory services, like the drug revolving fund. Doing the same for laboratory services would greatly aid us in purchasing reagents, equipment, maintenance, and other related costs” (**Government Hospital, Health Service Administrator**).*

#### Quality assurance

The study also examined compliance with essential quality assurance practices and procedures, such as laboratory accreditation and external quality assessment, necessary for delivering high-quality laboratory services.

##### External quality assessment

Many health managers consider external quality assessment (EQA) crucial for delivering high-quality laboratory services. However, it’s worrying that only laboratories within tertiary hospitals participate in EQA programs. The primary reason for this limited participation is the cost associated with these programs.

> *“The laboratory does not currently participate in External Quality Assessment Programs. However, as a manager and clinician, I believe that participating in these programs would enhance our services and provide reassurance to clinicians regarding the accuracy of our results.”* (**CHAG Hospital, Medical Director**).
>
> *“A national external quality assessment program for HIV and malaria has been inactive for some time, leaving laboratories without participation. The high costs of implementing quality assurance procedures are a significant barrier. To promote accuracy, we have advocated for regional peer sample transfers, but establishing a nationwide system remains costly”* (**Regional Director of Laboratory Services).**

##### Laboratory accreditation

Most laboratories in the region, except for one private lab, do not have accreditation to local or international standards. Participants noted that this is mainly due to a lack of information and the absence of a mandatory requirement for laboratory accreditation in Ghana.

> *“No public laboratory is accredited in this region; only one private laboratory claims to have been accredited to the ISO15189 standard. The absence of policy guidance or legal requirements for facilities to obtain accreditation is the primary obstacle to standardizing laboratories through accreditation”* (**Regional Director of Laboratory Services**).
>
> *“Although the lab is not currently accredited, accreditation is beneficial as it helps to ensure that the services are of high quality. Most labs lack accreditation because hospital managers are unaware of its importance” (**Private Hospital, Laboratory Manager**).*

#### Human resource capacity

The adequacy of laboratory personnel, training, and professional development emerged as the key components of human resources that influence the delivery of quality laboratory services.

##### Adequacy of laboratory personnel

Many laboratory managers believe that insufficient staffing and excessive workloads adversely impact the quality of laboratory services.

> *“Our round-the-clock services exacerbate the insufficient number of lab professionals at our facility. This situation adversely impacts the timeliness of results and extends patient waiting times. The shortage of staff is the primary reason for the prolonged turnaround time”* (**CHAG Hospital, Laboratory Manager**).
>
> *“The insufficient staff combined with a heavy workload causes long workdays and exhaustion, which can lead to errors and compromise the quality of laboratory services”* (**CHAG Hospital, Laboratory Manager**).

##### Training and continuous professional development

Most laboratory professionals report a lack of training programs unless they initiate them at their own expense.

> *“The facility lacks a structured training program for laboratory professionals. It’s been so long since I last attended any workshops. I believe that management should prioritize professional development and training to enhance the knowledge and skills of laboratory professionals”* (**Private Hospital, Laboratory Manager**).
>
> *“There is no system for training laboratory professionals in this facility until it is needed. I recall that some time ago, we organized training because an issue needed to be addressed, so we had to train all the staff at the front desk. We need to look at our training when it comes to laboratory quality management”* (**Private Laboratory, Laboratory Manager**).

#### Laboratory workspace and facilities

Most health managers’ report that the laboratory workspace does not optimize its operation and is a major barrier to providing quality laboratory services. It is interesting to note that, although the respondents are at the helm of affairs when it comes to decision-making regarding healthcare, it appears they are helpless in dealing with issues regarding laboratory services, as reflected in their comment:

> *“I do not think the current infrastructure facilitates optimal laboratory service provision because our greatest problem is space. This facility was an office complex converted into a hospital, so very few services were being offered from the beginning. So, the lab initially didn’t need much space, but now we have outgrown that initial situation. We are trying to do some expansion work, which is very capital-intensive. And we need all sorts of approval from stakeholders.” (**CHAG Hospital, Medical Director**).*
>
> “I believe the current infrastructure does not fully optimize our operations; however, we must adapt to the existing conditions. The facilities were not originally designed as laboratories, yet a hospital cannot function effectively without one. Ideally, we would have a single building housing a central lab, but I understand that this requires significant capital investment. As much as we desire this improvement for better service delivery, we must work with what we have for now” (**Tertiary Hospital, Director of Laboratory Services**).

#### Laboratory policy and regulation

Some health managers’ report that implementing laboratory policies and establishing a regulation system for laboratory services will enable laboratories to flourish, fulfil their mandate, and enhance the quality of medical laboratory services.

> *“A national laboratory policy, which addresses leadership and governance, quality assurance, human resources, infrastructure, laboratory space, and test menu per level, would enable quality service.”* (**Regional Director of Laboratory Services**)
>
> *“Laboratory regulatory bodies must enhance their efforts to ensure standardized services. The growth of private laboratories is concerning, as profit should not compromise patient care. These bodies must enforce standards to guarantee the quality of laboratory services.”* (**Metropolitan Director of Health Services**).

## Discussion

The analysis identified key barriers to delivering quality medical laboratory services, including laboratory financing, quality assurance, human resource capacity, laboratory workspace and facilities, and laboratory policy and regulation.

According to Travers (1996), “From a financial standpoint, one of the most valuable assets in the survival of a healthcare organization is the clinical laboratory”[18]. Laboratory financing and inadequate budget allocation have been recognized as major obstacles to providing quality medical laboratory services, especially in low- and middle-income countries [19,20]. This aligns with the findings of this study, where financial constraints in laboratory funding were identified as significant barriers to delivering high-quality medical laboratory services. Although the study participants may not fully grasp the complexities of financial management in healthcare, they acknowledge the importance of implementing sound financial practices to promote increased investment in medical laboratory services. Hermansen (2020) argued that reduced reimbursement and the need to control costs continue to place pressure on clinical laboratories [21]. Financial management becomes even more challenging due to shared revenue streams and interconnected cost structures when the clinical laboratory operates within a hospital setting [21]. Most participants emphasized this point in their comments, suggesting that while laboratories generate the highest revenue for the hospitals, this income is absorbed into the hospital’s overall finances. Evidence suggests that laboratory services account for less than 2.5% of hospital expenses but guarantee more than 300% profitability, making laboratory diagnostics a highly profitable business [22]. A laboratory that is financially and operationally successful is better positioned to support the broader goal of high-quality healthcare [21]. Most participants proposed establishing a separate laboratory fund for retooling medical laboratory services, though some expressed hesitancy from policymakers and health managers to implement such measures. Horton et al. (2018), in their assessment of laboratory services in low- and middle-income countries, suggested that diagnostic testing should be part of decision-making processes, supported financially, and that at least 4% of healthcare expenditure should be allocated for laboratory services to ensure high-quality and affordable testing [23].

Quality assurance encompasses all actions and steps undertaken by medical laboratories to enhance the standard and clinical value of test results. Laboratory services are highly technical and demand rigorous programs to ensure test results’ accuracy, reliability, and timeliness [10]. Notably, while all participants acknowledge the importance of external quality assessments and accreditation in guaranteeing the delivery of quality medical services, none of the laboratories evaluated, except for the teaching hospital, participate in external quality assessment programs or hold accreditation to ISO 1589 laboratory standards. External Quality Assessment (EQA) programs are designed to enable medical laboratories to compare the accuracy of their test results with those of their peers through statistical assessment against an expected or target value [24]. The accreditation program for medical laboratories has been shown to improve laboratory processes, enhance patient care, reinforce quality standards, reduce errors and risks, and lower test costs[25,26] A survey among laboratory professionals across Mediterranean countries assessing participation in PT schemes reveals that 53% participate because it is mandated by law, 29% follow scientific society guidelines, 6% due to social security claims, and 47% do not participate because it is voluntary [27]. These findings support the study’s results, indicating that the lack of engagement in external quality assessment (EQA) and accreditation by laboratories in Ghana stems from the voluntary nature of these processes. Previous research in other African countries also highlights that international accreditation is often prohibitively expensive, making it unaffordable for many low- and middle-income countries [28][29]. Quality does not come cheap, and decision-makers should understand that every dollar invested in quality yields savings through more efficient, prompt diagnostics and a reduction in patient harm[30]. The current cost constraints in laboratories hinder efforts to improve quality[31]. Indigenous accreditation programs could be pivotal in maintaining quality laboratory services in Ghana.

The laboratory personnel are the most vital resource and are essential to the provision of high-quality services. Many participants indicate that a shortage of laboratory staff impedes service delivery, a concern supported by numerous studies. Globally, a lack of workforce capacity significantly hampers access to quality laboratory services, and the recent COVID-19 pandemic brought to the fore the human resource challenge [4,32,33]. Some laboratory experts suggest that perhaps the most important lesson for policymakers and health managers during this pandemic is to invest in human resources, as an insufficient workforce and technical expertise can impact the management of emerging health crises[4,30]. The inadequate capacity of laboratory human resources is affecting healthcare systems worldwide, requiring coordinated efforts to address it. The rapid evolution of testing methods and instrumentation underscores the need for ongoing professional development to maintain clinical competence. However, most participants report lacking any formal training and development system, which is concerning because continuous professional development is crucial for delivering accurate, reliable, and timely laboratory services. Interestingly, similar observations have been made in other studies: most LMICs lack adequate capacity for ongoing skill enhancement and professional growth[10,34].

The medical laboratory is a workplace where various hazardous chemicals, complex instrumentation, and potential pathogens are encountered daily [35]. The laboratory’s work environment and facilities must be designed to ensure that tasks can be completed without compromising quality or endangering the safety of staff, patients, and the general public[36]. The study found that most hospitals were not designed for their purpose, leading to poor laboratory layouts and designs. This ultimately affects workflow and compromises the safety of laboratory staff, patients, and the broader community. Wilson et al. (2018), in their assessment of laboratory services in low and middle-income countries, identified inadequate infrastructure as a critical barrier to providing optimal laboratory services in the sub-region [10], confirming what was observed in this study. In Ethiopia, an assessment conducted in laboratories revealed that 91.7% of them had issues with layout and inadequate space, resulting from poor engineering and a lack of knowledge regarding laboratory construction requirements [37], confirming the study’s findings. Understanding the central role laboratory services play in a well-functioning healthcare system at the policy and governmental level is crucial to overcoming the laboratory infrastructure gap in Ghana.

Research has demonstrated that a primary challenge for national laboratory systems in resource-limited countries is the lack of clear policies and effective leadership[34,38], which ultimately affects the quality of laboratory services. Although Ghana has made several attempts to develop a health laboratory policy and strategic plan, the process has stalled. Participants in the study believe that proper laboratory policies and regulation of laboratory services will create an environment where laboratories can thrive and improve the quality of medical laboratory services. The Maputo Declaration on Strengthening Laboratory Systems encourages governments to develop national laboratory policies as part of their national health development plans [39]. The National Laboratory Policy sets minimum standards for laboratory services necessary to support essential clinical and public health services at all levels of the healthcare system. It also addresses organizational and management structures, regulatory mechanisms, human resource needs, and all necessary support services within the healthcare delivery system[40]. The absence of a national laboratory policy and its implementation led to fragmentation of laboratory services, affecting the provision of quality services, and resulting in suboptimal health outcomes.

## Conclusion

Barriers to providing quality medical laboratory services include issues related to laboratory financing, quality assurance, human resource capacity, and policy and regulation. To address these challenges, implementing a comprehensive national health laboratory policy is essential. This policy will create a framework for the coordinated development and delivery of accessible, high-quality laboratory services throughout Ghana.

## Data Availability

Qualitative data was collected from human subjects and cannot be publicly shared. The data is available from the corresponding author ES at upon request.

## Acknowledgement

We would like to express our gratitude to the research team, all hospitals, Health Directors, Health Service Administrators, Medical Superintendents, Nurse Managers, and Laboratory Managers who assisted with recruiting study participants and data collection.

